# Exploring the utility of dynamic motor control to assess recovery following pediatric traumatic brain injury: A pilot study

**DOI:** 10.1101/2025.04.17.25324814

**Authors:** Nanette Aldahondo, Andrew J. Ries, Amy Schulz, Michael H. Schwartz

## Abstract

**Background:** Pediatric traumatic brain injury can lead to severe disability. Currently used standard clinical measures effectively capture secondary functional impairments, but do not measure neurologic impairment directly.

**Objective:** Evaluate the feasibility of walking dynamic motor control (walk-DMC) assessments to more directly measure neurological impairment and recovery for individuals post-traumatic brain injury.

**Methods:** The trajectory of walk-DMC and standard clinical measures of balance, mobility, and function were assessed in a cohort of individuals post-traumatic brain injury. Measures were collected throughout participants’ inpatient rehabilitation stay and at short- and long-term follow-up assessments.

**Results:** Four pediatric participants with severe traumatic brain injury enrolled. All participants demonstrated substantial neurological impairment at enrollment. All clinical measures showed an initial deficit followed by recovery, with most returning to nondisabled ranges over the study period for all participants. In contrast, walk-DMC scores demonstrated an initial acute deficit and did not reach nondisabled ranges for two of the participants, indicating persistent neurologic impairment.

**Conclusion:** Walk-DMC shows promise in its ability to identify subtle ongoing neurologic impairment compared to traditionally used clinical assessments of balance, mobility, and function. Further work in a larger cohort of participants with traumatic brain injury will improve understanding of how walking dynamic motor control changes with injury severity and where such a measure can serve as a leading indicator of neurologic and functional recovery.

## Introduction

Traumatic brain injury (TBI) in children is a significant public health burden in the United States and can lead to severe disability. It has been estimated that more than 60% of children with moderate to severe TBI experience long-term disability(1), with the majority having complete or near-complete loss of walking ability in the acute period(2) and persistent functional deficits even 36 months post-TBI(3–5). For families, recovery of walking ability is often a central concern due to its importance for independence, and most pose questions to their child’s physician such as ‘Will my child walk’, ‘If so, when will they walk?’, and ‘How well will they walk?’.

Because TBI is a heterogeneous injury, answering these types of questions is often challenging. The extent of movement impairment is related to the location, nature, and severity of the brain injury. In the pediatric population, predicting recovery of function is further complicated by the varied developmental stages of each patient at the time of injury(6) as well as other issues such as patient demographics (e.g. age), and hospital factors (e.g. initial Glasgow Coma Scale score, presence/length of coma, length of hospitalization)(7,8). This heterogeneity in recovery trajectory is reflected in the current literature. Studies examining functional recovery after pediatric TBI have described improvement from six months post-injury up to two years after injury, with some children showing ongoing functional gains even after two years(7,9,10). Studies have also shown that functional recovery slows over time and children may have continued functional deficits as compared to non-injured matched peers three years post-TBI(3–5,9,11).

As highlighted in a rehabilitation engineer’s frank and impassioned commentary on his young child’s TBI, ‘Unfortunately, there is a dearth of clinical evidence that shows clinicians and parents what treatments and strategies are most beneficial.’(12) Indeed, many current treatment paradigms consider the brain as a black box and, therefore, often rely on a ‘wait and see’ approach to recovery. Clinicians treating children after a severe TBI and their families have a strong motivation to prognosticate recovery as early as possible to inform treatment and guide recovery expectations. Unfortunately, the substantial variability in recovery from a TBI only allows clinicians to offer vague answers about recovery which can leave patients and families frustrated and unsatisfied. This highlights a significant gap in clinical care—there is currently no measure that can help clinicians reliably address questions regarding recovery of function after TBI.

The variability that has been previously reported in TBI recovery may be due, in part, to the limitations of current assessment methods employed to monitor various aspects of balance, mobility, and functional recovery. The current literature is largely based on clinical measures of recovery that, although well accepted, assess neurologic impairment indirectly via functional tests of balance and mobility and may not be sufficiently discriminating to identify subtle but ongoing deficits higher up in the motor control hierarchy(9–11,13). Additionally, these measures are lagging indicators of recovery meaning that they can capture a person’s current abilities and their compensations and improvement to date but cannot provide prognostic value for future abilities/recovery. As such, identifying alternative discriminating methods that can assess acute functional deficits and longitudinal recovery, and simultaneously serve as a leading indicator of future recovery is clinically and socially relevant to help guide treatment and frame expectations. Ideally, these alternative methods would lead to improved assessment of the neurological injury and recovery tracking and generate better-informed treatments to supplement and enhance the recovery process. Walking dynamic motor control (walk-DMC) is a measure of motor control complexity derived from muscle activity data collected during walking(14). The foundational assumption for walk-DMC is that the brain partially controls cyclic movement through a series of synergies or motor modules(15). These synergies are patterned muscle activations that simplify the brain’s motor control demands during cyclic tasks. Walk-DMC has been used to evaluate neuromuscular function for individuals with static neurologic injury, such as those diagnosed with cerebral palsy (CP). Earlier work in CP has not only demonstrated that walk-DMC is reduced compared to nondisabled peers but has a direct effect on functional gait outcomes including Gross Motor Function Measure (GMFM) scores, walking speed, energy expenditure, and self-reports of community mobility(14,16–18). Previously, a series of studies utilized walk-DMC to assess the complexity of neuromuscular control in adults with chronic mild to moderate TBI(19,20). These studies found that adults with chronic TBI had decreased neuromuscular control complexity (i.e., lower walk-DMC scores) compared to age-matched nondisabled peers. While evidence is still limited in TBI, this work suggests that walk-DMC may be a useful metric for tracking meaningful changes in neuromuscular function and prognosticating recovery among children post-TBI(19,20). However, walk-DMC has yet to be evaluated in this context.

The goal of this study was to assess the utility of walk-DMC to quantify the severity of the initial neurologic injury and measure changes in impairment throughout the recovery process in pediatric patients post-TBI. We measured walk-DMC alongside other typically used assessments of balance, mobility, and function in a cohort of patients with acute TBI over the course of their inpatient rehabilitation stay as well as at short-and long-term follow-up assessments. We hypothesized that walk-DMC would be reduced in acute TBI compared to typically developing peers, reflecting a simplified motor control strategy, similar to prior studies in CP (14). Furthermore, we hypothesized that walk-DMC would change at a different rate than standard outcome measures, demonstrating discriminative value.

## Material and Methods

### Participants

This was an exploratory study with a convenience sample of children and adolescents (ages 4-18) admitted to an inpatient rehabilitation unit after sustaining a severe TBI. All study procedures were approved by the University of Minnesota Institutional Review Board (#00006147). Written informed consent was obtained from the participant’s legal guardian and verbal or written assent was obtained as appropriate in participants 8 years or older. Individuals admitted to the inpatient rehabilitation unit post-TBI were screened for this study. Recruitment was performed between January 2020 and December 2020 through distribution of an introductory letter explaining the study after individuals were screened as eligible by the principal investigator. To participate, individuals needed to have an initial Glasgow Coma Scale (GCS) ≤8 after TBI indicating severe neurological injury, be able to walk approximately 30 steps regardless of level of support/assistance at the time of assessment and be able to cooperate with testing procedures. Individuals were excluded if they had a history of prior developmental delay, preexisting motor impairment, progressive neurologic disease, or recent injury other than the TBI which affected their ability to ambulate (e.g., lower extremity fracture, peripheral nerve injury). The GCS was collected as part of the standard of care immediately after TBI and was only used to determine study eligibility.

### Assessment Protocol

As soon as participants were able to walk 30 steps, the study team began obtaining twice-weekly assessments for the first two weeks, followed by once-weekly assessments until discharge from the inpatient rehabilitation unit. Short-term and optional long-term follow-up data were collected at 6-months and 2-years post-TBI, respectively. Measures of balance, motor control, and functional ambulation were collected during each assessment.

At each assessment, the study therapist placed electromyography (EMG) electrodes according to SENIAM guidelines(21) on the anterior tibialis, medial gastrocnemius, vastus lateralis, rectus femoris, and medial hamstrings of the participant’s most severely affected leg. Participants then completed the Pediatric Balance Scale (PBS) to assess balance and the Timed Up-and-Go (TUG) test and 6-minute walk test (6MWT) to assess functional gait performance. The study therapist provided the minimum amount of assistance needed for the participant to complete each clinical test and documented this value according to WeeFim® guidelines using a 1-7 grading scale (1-Total Assistance, 7-Complete Independence). Participants did not wear their orthotics while completing the 6MWT. In addition to these standard clinical measures, EMG data collected during the 6MWT was used to calculate walk-DMC. A robust walk-DMC estimate for 6-minutes of EMG data was derived using 10 random 60-second bouts of muscle activity data sampled from the full 6-minute trial. A walk-DMC value was calculated—as previously described(14)—for each of the 10 bouts and the median value was used as the overall walk-DMC score for the assessment session. The 60-second time was chosen to include data from a sufficient number of gait cycles to allow for a stable walk-DMC calculation(22).

Throughout recovery, measures of community function were also collected. The Community Balance and Mobility Scale (CB&M) was completed, as able, at the final assessment before discharge and at both the short- and long-term follow-up assessments. The CB&M captures dynamic balance and mobility deficits based on tasks commonly encountered in the community in higher functioning ambulatory individuals with TBI(23,24). The items tested are more challenging than those tested in other measures and require speed, precision, and recovery of stability after voluntary perturbations. In addition, the CB&M has not shown a ceiling effect like other measures of balance and mobility(23–25). Additionally, the Child and Family Follow-up Survey (CFFS) was completed at the short-term follow-up and a self-report questionnaire (included in Appendix) was completed at the long-term evaluation to obtain a qualitative assessment of the participant’s recovery. The CFFS measures outcomes and the needs of children with TBI across several domains with subscales that look specifically at participation in the home, school, and community settings (The Child and Adolescent Scale of Participation; CASP), injury related factors such as health, cognitive, psychological, physical, and sensory functioning (The Child and Adolescent Factors Inventory; CAFI), and environmental factors related to barriers in the home, school, and community settings (The Child and Adolescent Scale of Environment; CASE). Caregivers also rated the participants current mental and physical health as well as their current quality of life (QOL)(26). Finally, a self-report questionnaire (S1 Appendix) was created by the investigators to document perceived recovery in mobility, memory, and mood functioning. To capture each individualized perspective, both participants and their caregivers were asked to rate their current mobility, memory, and mood using a Likert scale ranging from ‘a lot worse than before the injury’ to ‘a lot better than before the injury’. A summary of all standard and experimental measures collected during this study is provided in Table 1.

**Table 1.** Outcome measures summary.

| Domain | Test | Areas Assessed | When Assessed |
| --- | --- | --- | --- |
| ICF Body Function | Pediatric Berg Balance Scale (PBS) | Functional balance (static and anticipatory) | Inpatient Period: Twice weekly for the initial 2 weeks of the study, then weekly until discharge<br>Post-Discharge: Short-Term (6 months-post injury) and Long-Term (~ 2 years post-injury) Follow-Up |
|  | Timed-up-and-go test (TUG) | Balance, functional mobility | Inpatient Period: Twice weekly for the initial 2 weeks of the study, then weekly until discharge,<br>Post-Discharge: : Short-Term (6 months-post injury) and Long-Term (~ 2 years post-injury) Follow-Up |
|  | 6-minute walk test (6MWT) | Functional walking capacity | Inpatient Period: Twice weekly for the initial 2 weeks of the study, then weekly until discharge<br>Post-Discharge: : Short-Term (6 months-post injury) and Long-Term (~ 2 years post-injury) Follow-Up |
|  | Community Balance and Mobility Scale (CB&M) | High level balance and mobility | Inpatient Period: As able at discharge<br>Post-Discharge: : Short-Term (6 months-post injury) and Long-Term (~ 2 years post-injury) Follow-Up |
| ICF Body Structure | Walk-DMC | Neurologic impairment | Inpatient Period: Twice weekly for the initial 2 weeks of the study, then weekly until discharge<br>Post-Discharge: Short-Term (6 months-post injury) and Long-Term (~ 2 years post-injury) Follow-Up |
| ICF Activity and Participation | Child and Family Follow Up Survey (CFFS) | Caregiver rating of physical and emotional well-being, participation, problems experienced in daily life, the impact of environmental factors, and quality of life | Post-Discharge: Short-Term Follow-Up (6 months-post injury) |
|  | Self-report questionnaire | Participant and Caregiver rating of mobility, memory, and mood | Post-Discharge: Long-Term Follow-Up (~ 2 years post-injury) |

### Statistical Analysis

Due to the small sample size of this study, statistical analysis is largely descriptive. Clinical outcome scores were converted to z-scores—where applicable—using age and sex-based normative values in order to standardize outcome comparisons. (walk-DMC(14), PBS(27), 6MWT(28,29), TUG(30), CB&M(25,31)).

## Results

### Study Sample

Four participants with severe TBI were enrolled in this pilot study (Table 2). The principal investigator, study therapist, and study staff determined the more impaired side according to clinical measures of strength and motor control.

**Table 2.** Participant characteristics.

| Participant | Age (y) | Sex | Mechanism of injury | Initial GCS | Days from injury to inpatient rehab | Days from injury to enrollment | Days from injury to hospital discharge | Impairment Laterality |
| --- | --- | --- | --- | --- | --- | --- | --- | --- |
| 1 | 5-10 | F | Generalized | 4 | 10 | 14 | 28 | Symmetric |
| 2 | 15-20 | M | Focal | 3T | 12 | 20 | 49 | Symmetric |
| 3 | 15-20 | F | Generalized | 3 | 15 | 78 | 100 | Right |
| 4 | 15-20 | M | Generalized | 3 | 18 | 38 | 83 | Left |

Despite similarities in initial GCS scores - indicating all participants had severe injuries - the timing of study enrollment and length of stay was variable across participants. Both Participants 1 and 2 were enrolled within a week of admission to the inpatient rehabilitation unit and had shorter lengths of stay, whereas participants 3 and 4 had a larger gap between admission to the unit and study enrollment and had at least double the length of stay as Participants 1 and 2.

All participants completed all assessments while in the inpatient rehabilitation unit. Due to COVID-19 restrictions at the time, Participant 1 did not complete either the short- or long-term assessment and Participant 2 did not complete the long-term assessment. Participant 1 was able to complete the CB&M prior to discharge while all other participants completed the CB&M at the short-term follow-up visit. Participants 3 and 4 completed both short- and long-term assessments.

### Functional Measures

At study enrollment all participants demonstrated substantial deficits in standard clinical measures (PBS, 6MWT, TUG) as well as walk-DMC, however the rate of recovery differed across outcome measures (Fig 1). Standard clinical measures of function demonstrated consistent improvements over the study period, with all but the 6MWT returning to normal ranges by the short-term follow-up assessment. In comparison, walk-DMC scores showed some improvement over the course of the study, but appeared to recover at a slower rate than the clinical measures, and did not normalize for one of the participants even at the long-term follow-up assessment.

**Fig 1.** Longitudinal results post-TBI as measured for Walk-DMC, PBS, TUG, 6MWT, CB&M, and required walking assistance level. CB&M items reflect balance and mobility skills needed for full participation in the community. Higher scores indicate better balance and mobility. WeeFim® levels of independence were rated during the 6MWT. Note: x-axis is broken into two different time scales.

### Community Function and Follow-Up Surveys

Overall, Participants 1 and 2 had quicker recoveries in all objective outcome measures compared to Participants 3 and 4. This trend is also reflected in the CB&M scores, where Participant 1 was able to complete this test prior to discharge, reflecting their level of recovery, and both Participants 1 and 2 had markedly higher scores at the short-term assessment compared to Participants 3 and 4 (Figure 1). Participants 3 and 4 did have some improvement in CB&M scores between short- and long-term assessments but still had suppressed scores at the long-term assessment, indicating a significant level of remaining impairment.

The caregivers of Participants 2, 3, and 4 completed the CFFS during the short-term follow-up assessment (Table 3). Overall, caregivers rated the participant’s physical and emotional health and well-being as good to excellent. Similarly, QOL for all participants was rated as excellent. However, the subscores for participation (CASP), injury limitations (CAFI), and environmental barriers (CASE) indicate remaining challenges. Interestingly, there was a large difference in participation scores between Participant 2 and 4 despite both caregivers reporting nearly identical scores for injury limitations and environmental barriers.

**Table 3.** Child and Family Follow-up Survey (CFFS) results.

| Participant | Physical health | Emotional health and well-being | Participation (CASP) | Injury Limitations (CAFI) | Environmental Barriers (CASE) | QOL |
| --- | --- | --- | --- | --- | --- | --- |
| 1 | - | - | - | - | - | - |
| 2 | Good | Good | 25 | 40 | 35 | Excellent |
| 3 | Excellent | Excellent | 46 | 56 | 48 | Excellent |
| 4 | Very Good | Excellent | 94 | 38 | 35 | Excellent |

Scores conformed to a 100-point scale. CASP: higher numbers indicate greater participation. CAFI: higher numbers indicate greater level of impairment/greater extent of problem. CASE: higher numbers indicate greater extent of environmental barriers. Participant 1 did not complete the short-
term follow-up assessment due to COVID-19 restrictions. Surveys were collected at the short-term follow up.

At the long-term follow-up assessment, both Participants 3 and 4 and their caregivers differed in their perceptions about recovery of mobility, memory, and mood (Table 4). On the self-report questionnaire, both participants described their own mobility as being ‘a little worse than before the injury’. In contrast, the caregivers for Participants 3 and 4 described mobility as being ‘a lot worse than bef ore the injury’ and ‘not better or worse than before the injury’, respectively. Perceptions about memory and mood recovery varied between participants and caregivers as well.

**Table 4.** Self-report questionnaire.

| Participant | Mobility |  | Memory |  | Mood |  |
| --- | --- | --- | --- | --- | --- | --- |
|  | Self | Caregiver | Self | Caregiver | Self | Caregiver |
| 3 | Little worse | Lot worse | Little worse | Lot worse | Not better or worse | Little better |
| 4 | Little worse | Not better or worse | Not better or worse | Little worse | Little worse | Little better |

## Discussion

This pilot study indicates that using only standard clinical assessments to monitor recovery from TBI may mask ongoing functional deficits, leading to a significant discrepancy between perceived and measured outcomes. These clinical measures are lagging indicators of recovery and do not capture the compensatory strategies a person may use to overcome continued neurologic impairment. In contrast, measures of walk-DMC provide evidence of persistent impairments to neurologic function which is consistent with clinician and participant/caregiver reported impressions. These results highlight the importance of examining neurological impairment as directly as possible and supports further investigations of dynamic motor control as a potential tool for monitoring acute neurologic impairment and functional recovery in children and adolescents after severe TBI.

Our findings suggest that for individuals with TBI, walk-DMC has the potential to capture salient features related to neurologic impairment that are not reflected by standard clinical measures of balance, mobility, and function post-TBI. In line with our original hypothesis, this study demonstrated that walk-DMC was reduced following severe TBI and remained depressed at short- and long-term follow-up assessments in contrast to the standard clinical measures of balance, mobility, and function. This was most noticeable when examining Participant 3’s recovery trajectory. The persistent deficit in walk-DMC observed indicates that they continued to utilize a simplified motor control strategy at the time of inpatient discharge, similar to what has been observed for individuals with CP and adults with chronic TBI(14,19,20).

The results of this study are intriguing and warrant further investigation. Futures studies should strive to answer questions such as: Is walk-DMC a reliable measure for tracking neurologic impairment? Does it vary according to injury severity? Can it serve as a leading indicator of neurologic recovery, and does it offer enough specificity to add prognosticative value? If so, will it help inform beneficial interventions?

### Study limitations

There are several limitations and challenges to consider when interpreting these pilot data. First, we noted a significant amount of variability in walk-DMC values between assessments, making it difficult to separate true change in motor control from experimental noise. Analysis of the walk-DMC calculation methodology (i.e. using the median walk-DMC derived from 10 60-second EMG samples) indicates that the intra-session walk-DMC calculation was quite reliable (the maximum walk-DMC range for any single participant assessment across the 10 samples was 3.6 points), and that a single random 60-second bout of EMG data may be sufficient to provide as reliable a walk-DMC measurement as from an analysis of the full 6-minutes of EMG data. However, we suspected that the main factor influencing the reliability of walk-DMC in this study is uncertainty regarding the repeatability of electrode placement (i.e. precise and appropriate muscle identification) between assessments (i.e. inter-session variability). As such, differences in walk-DMC scores observed between assessments may not only be caused by changes in motor control but may also be affected by experimental error. Previous studies of individuals with CP (i.e. individuals with static neurologic impairment) have shown that the inter-session reliability of walk-DMC is approximately 6 points(14,32). However, compared to these previous studies, our study team was relatively inexperienced at the early data collection phases regarding EMG electrode placement. As such, we anticipate that a significant portion of the variability in walk-DMC scores measured during the course of this study was due to experimental error caused by variability in EMG sensor placement between assessments (i.e. cross-talk, erroneous muscle activity), plausibly leading to inter-session variability greater than 6 points. While it is unclear how inter-session walk-DMC reliability translates between CP and TBI patient populations, gaining additional training and experience as well as developing robust methods to assist in the consistent placement of electrodes (e.g. temporary markings) would improve the reliability of measured walk-DMC no matter the sample population.

In addition to the variability in measured walk-DMC values, TBI is a heterogeneous injury which makes it challenging to draw definitive conclusions from the limited number of participants in this study. The acuteness of the injury also poses additional challenges. Families are often just coming to terms with the injury and starting to grieve when they transfer from the acute care hospital to inpatient rehabilitation, which can make it difficult to standardize recruitment and timing of enrollment. Furthermore, to participate in this study, individuals needed to be able to ambulate, which, depending on the severity and location of the injury, may results in a significant delay between the injury and study enrollment. In this cohort, despite all participants having severe neurologic injury, as described by initial GCS, their mechanisms of injury varied significantly, as some had a more diffuse neurologic injury pattern than others. Other surrogate measures of severity such as the time from injury to inpatient rehabilitation and lengths of stay were also variable. In addition, all participants’ initial levels of function and recovery of function were substantially different from one another, meaning that the level of support required for walking trials differed significantly between individuals and over time. For example, during the initial assessment, one participant required bodyweight support provided by a TRAM and the assistance of two therapists, while another only required a gait belt. Although, studies have shown that synergies are unaffected at different levels of purely vertical support or different conditions of walking, the extent of compensation, offloading, and assistance required differed at different times throughout the study period and likely affected measurements. (33,34)

Additionally, it is clear that 6MWT outcomes did not normalize for any of the participants, as the majority were still more than three standard deviations from normal at long-term follow-up. While these results may indicate persistence functional deficits, there may be alternative explanations for the abnormal results observed here. First, systematic differences in instructions and participant interpretation during the study may lead to lower 6MWT scores. Alternatively, cardiovascular fitness may have been impaired due to the dramatic reduction in physical activity experienced by the participants post-TBI, in which case, the applicability of the 6MWT as a method for assessing neurologic impairment and recovery after a significant decrease in physical activity may be questioned. Future studies should work to examine if these alternative explanations have any impact on the validity of 6MWT for TBI.

Finally, while walk-DMC may be an objective and clinically valuable measure of motor control in ambulatory patients with acute TBI, challenges like the variable timeline of recovery and the need for varying levels of assistance (especially during the initial stages of recovery) bring into question the feasibility and reliability of using walking as the predominant task to assess dynamic motor control, especially for the acutely injured. Because walking requires complex integration between many functions (balance, muscle coordination, visual perception, limb proprioception, etc.) it is inherently variable over the course of recovery from neurologic injury, and the interplay between these functions likely impacts data reliability. This points to the potential need for an alternative activity that can be used to quantify dynamic motor control through all stages of recovery, including the acute phase. Recumbent pedaling may be a promising alternative, as it is commonly used as an early pre-ambulation rehabilitation strategy in patients after neurologic injury. Additionally, pedaling and walking share similar complex coordinated reciprocal movements (e.g. reciprocal flexion/extension movements, alternating activation of antagonist muscles) but pedaling an adaptive or stationary bicycle requires less balance than walking and can be more easily controlled experimentally, improving the consistency of data collection. As such, exploring pedaling as a means of quantifying motor control throughout all stages of recovery post-TBI and its utility as a leading indicator of potential neurologic recovery is an exciting avenue for future studies. This concept of shared synergies in cycling and walking is already being investigated. (35,36)

## Conclusions

Pediatric TBI remains a significant public health burden in the United States and can lead to severe disability. Identifying a quantitative metric to measure impairment severity, track functional recovery, and potentially serve as a leading indicator to guide intervention is a key goal for improving treatment and enhancing recovery post-TBI. This study demonstrated that a quantitative assessment of motor control, walk-DMC, may be able to identify ongoing neurologic impairment and serve as a complement to standard clinical outcome measures. However, the small sample size and heterogeneity of injuries and recovery in this study makes it difficult to draw firm conclusions about the value of walk-DMC for assessments of TBI. Further work is needed to understand how walk-DMC and other complimentary quantitative measures of motor control are affected by injury severity, how they change throughout the course of recovery, and whether they add prognostic value.

## Data Availability

All relevant data are in the manuscript and supporting information files

## Acknowledgements

The authors thank Alyssa Spomer, PhD, Stacy Ngwesse, CCRC, and Elizabeth Nelson, MPH, CCRC for their valuable assistance in conducting this work.

